# FoodScribe: an open-source semantic framework for nutrient estimation from free-text dietary records

**DOI:** 10.64898/2026.07.15.26358181

**Authors:** Harsha Gouda, Marta Sala-Climent, Julius Agongo, Sayli P. Gaikwad, Annie Nattakom, Haoqi Nina Zhao, Shipei Xing, Brigid S. Boland, Tiffany Holt, Monica Guma, Pieter C Dorrestein

**Author notes:** **Corresponding Author** Pieter C. Dorrestein − Skaggs School of Pharmacy and Pharmaceutical Sciences and Collaborative Mass Spectrometry Innovation Center, University of California San Diego, La Jolla, California 92093, United States.

## Abstract

Efficiently summarizing dietary records at scale remains a persistent bottleneck in nutritional epidemiology. We present *FoodScribe*, which translates free-text meal descriptions into quantitative nutrient profiles by combining ingredient parsing with nutrient retrieval by querying the USDA FoodData Central (FDC) database. Benchmarked using three LLM providers using Nutribench dataset, FoodScribe completed annotation of 3,807 meal descriptions in 2.5 hours, a task otherwise requiring substantial manual effort from trained nutritionists. FoodScribe achieved accuracy across macronutrient estimation (F1=0.79-0.89), with models performing better for protein than fat estimation. Application to a Mediterranean diet intervention cohort indicated dietary shifts consistent with the intervention pattern based on model-derived estimates. Integration with metabolomics data suggested that fiber and vegetable intake were positively associated with a fecal metabolite cluster.

## Main

Dietary habits represent one of the most modifiable determinants of human health, influencing metabolic phenotype, microbiome composition, and disease risk^1–3^. Characterizing nutrient intake accurately is foundational to epidemiological research, precision nutrition trials, and mechanistic studies linking diet to chronic diseases^4–6^. Yet translating real-world dietary records into structured, quantitative nutrient profiles remains a time-intensive process, requiring trained nutritionists to interpret food descriptions, estimate portion sizes, and match items to standardized food composition databases such as the USDA Food Data Central (FDC)^7^. This process introduces variability through subjective interpretation, particularly for mixed dishes, recipes, or culturally diverse foods that are underrepresented.

In recent years, computational and technological approaches have emerged for dietary assessment^5,8–12^. Particularly, Image-based dietary assessment tools, barcode scanning applications using mobile apps have shown promising approaches to capture dietary intake reducing the burden on the participants^13^. Each method have inherent strengths and limitations. Diet logging in real time has shown promising results in improving health outcomes, as the food frequency questionnaires (FFQs) or 24-hr recall dietary approaches are prone to underreporting, as they rely on memory and self-estimation^14^. Dietary logs are collected on paper or digitally by having individuals record all food, beverages and supplements consumed in real time, where participant notes describe dietary intake with as much detail as possible with brand names, preparation methods with estimated portion sizes^15^. Then the food intake is linked to nutritional profile by leveraging the food composition databases such as FDC. The complexity of natural language for dietary reports is substantial, where vocabulary and language used to describe food is non-standardized and defined by the individual culture, schooling, training and environment. Example entries like “A plate of vegetable biryani with a side of raita, and a glass of mango lassi.” can refer to a meal with multiple ingredients with different portion sizes. Dietitian decoding of such meal descriptions can take 5-10 minutes based on the familiarity, to estimate portion size and look for appropriate food database identifiers. This constraint limits the scalability of dietary assessment in large cohort studies, where expert coding or third-party annotation services introduce substantial per-participant costs^16^.

The emerging technologies like the large language models (LLMs) are trained on an immense amount of text data from the web, including the reports of recipes, food composition tables, and nutrition-relevant literature^17,18^. Therefore, they can decode associations between colloquial food descriptions, their constituent ingredients, and plausible portion sizes, precisely the knowledge a dietitian applies when decoding a free-text meal entry, operating consistently and traceably at scale across thousands of records. Leveraging this capability, we developed FoodScribe, a two-stage computational pipeline that converts unstructured free-text meal descriptions into structured nutrient profiles grounded in the FDC database. In Stage 1, an LLM-based parses a structured list of ingredients and estimated gram quantities of each ingredient in a meal description (**Fig. 1a**). In Stage 2, FoodScribe performs retrieval using sentence-transformer embeddings to match each ingredient to its closest FDC database entry, from which nutrient values are scaled by predicted quantity and summed to yield meal-level nutrient totals (**Fig. 1a**). Unlike text matching, the semantic retrieval using sentence transformer embeddings maps food terms in close geometric proximity based on meaning, for example, an entry with “rocket leaves” will be matched to “arugula” despite no lexical overlap. Existing approaches rely on exact keyword matching, proprietary software, or closed commercial platforms. FoodScribe provides an open-source, transparent, and provider-agnostic framework that preserves traceability across all annotation steps. Prior applications of LLM models in dietary assessment often relied on the models to directly estimate macronutrients or output database identifiers, which can lead to untraceable errors and hallucinations. By restricting the LLM to parsing ingredients and using a separate sentence-transformer model for semantic matching to the USDA database, this approach ensures that all nutrient calculation is grounded in verified, standardized data.

**Figure 1:**
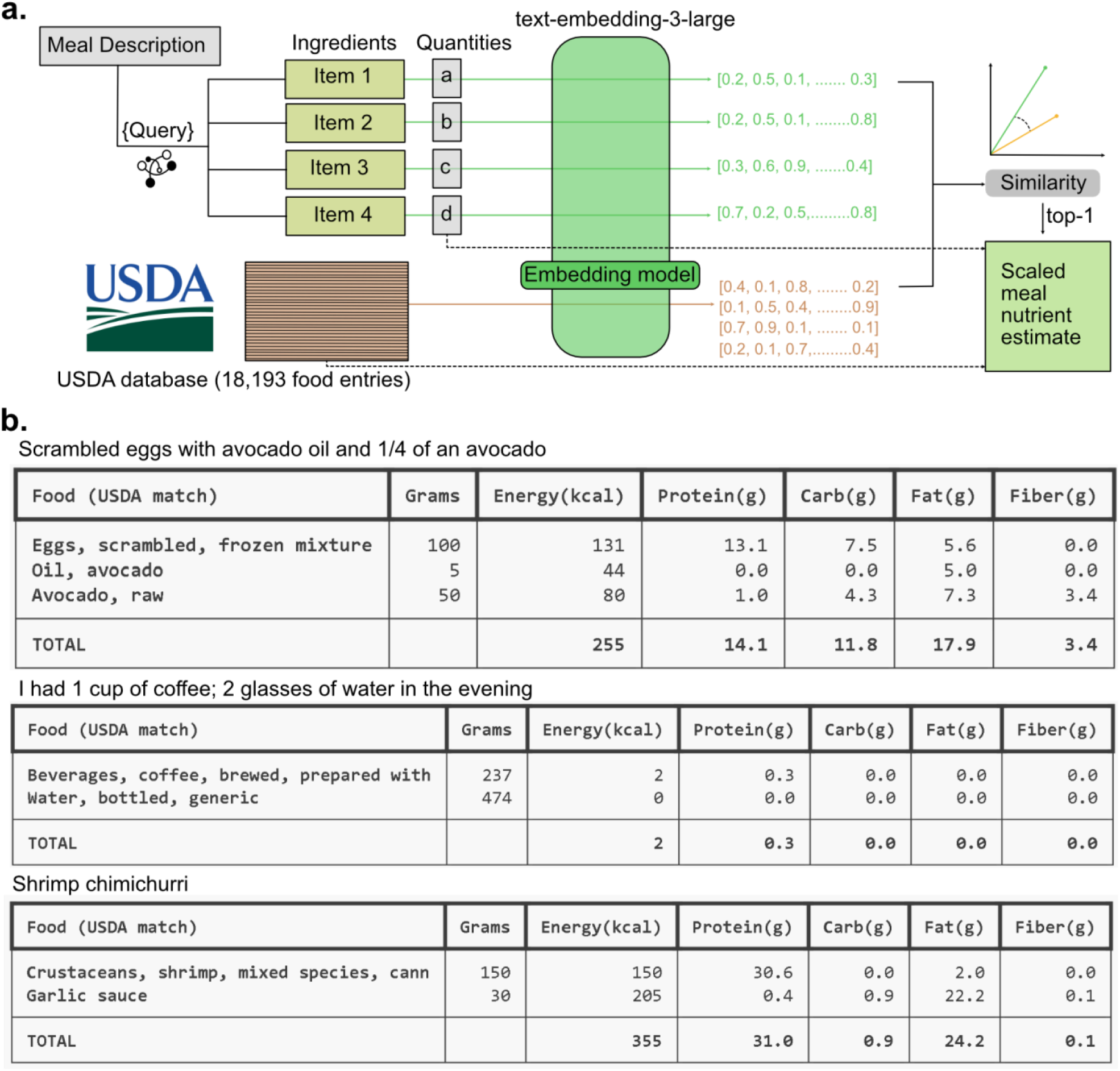
The FoodScribe workflow and examples. (a) large language model-based extraction of ingredients from natural language meal descriptions are used for semantic retrieval against the USDA FoodData Central entries, for dietary ingredient retrieval. (b) Example foodscribe outputs from representative meal entries from dietary records.

We benchmarked FoodScribe nutrient estimation using NutriBench-curated meal descriptions^19^. Meal descriptions from 16 countries (n=4,881) were analyzed using three LLM providers: DeepSeek (deepseek-chat, V3.2), OpenAI (GPT-5-mini), and Anthropic (Claude Sonnet 4.6). Performance was assessed for total calories (kcal), carbohydrates (g), protein (g), fat (g), and fiber (g) using mean absolute error (MAE) and F1 statistics. Model accuracy varied by geographic origin of meal descriptions, with higher performance observed for Bulgarian, Peruvian, Indian, and US-based meals compared to other countries, suggesting that cultural specificity of dietary descriptions influences estimation accuracy (**Fig. S1a**).

We then focused on the US-based meals (n=1,995) as they are most relevant to the USDA Food central database, OpenAI achieved the highest caloric accuracy (MAE=70 kcal), outperforming Anthropic (MAE=101 kcal) and DeepSeek (MAE=181 kcal). All three models exhibited a systematic overestimation bias, most pronounced for DeepSeek (+109 kcal) relative to OpenAI (+24 kcal) and Anthropic (+37 kcal). At a ±100 kcal tolerance threshold, F1 scores for caloric estimation were 0.88, 0.85, and 0.76 for OpenAI, Anthropic, and DeepSeek, respectively (**Fig. S1b**). Performance varied meaningfully by reporting style. Descriptions using metric quantities (e.g., “coffee with a teaspoon of sugar”) consistently outperformed natural language descriptions (e.g., “coffee with sugar”). For OpenAI, metric reporting yielded an MAE of 52 kcal, F1 of 0.922 (±100 kcal tolerance), and a bias of +19.2 kcal, with similar gains observed in carbohydrate, protein, fat and fiber estimation for the tested models. FoodScribe handled reporting style through a two-path strategy. When explicit quantity indicators were present, such as “2 cups of water,” “a slice of watermelon,” or “2 boiled eggs”. Reported quantities were extracted and converted directly to gram units for nutrient estimation. When no quantity was specified, FoodScribe defaulted to a standard single-serving size, based on predefined standard serving sizes (e.g., “I had macaroni and cheese for lunch” was interpreted as one standard serving). Estimating portion sizes from free-text descriptions is a time-consuming bottleneck in nutritional epidemiology, as users frequently omit exact measurements. The proposed methodology addresses this by automatically extracting metric quantities when present and systematically applying predefined standard serving sizes when explicit quantities are absent. This standardizes the handling of ambiguous entries at scale without requiring nutritionist intervention.

Following benchmarking, we applied FoodScribe for nutreint prediction from dietary records collected as part of a Mediterranean diet intervention study at the University of California San Diego (n=46; **Fig. 2a**). In a mediterranean (MED) dietary intervention trial, participants completed seven-day habitual diet logs followed by 14-day mediterranean diet diaries, yielding 3,807 free-text meal descriptions from 46 individuals (**Fig 2a**). FoodScribe parsed these entries into structured macronutrient, micronutrient and food group profiles in under three hours extracting over 230 nutrient variables as part of both habitual and mediterranean diet diaries. FoodScribe showed a reproducible and similar performance when the queried repeatedly (n=3) across the same model (**Fig. S2a-b**). Nutrient estimates derived from FoodScribe reflected differences in macronutrient composition consistent with Mediterranean dietary intervention. Following the MED diet intervention, caloric intake showed reduced inter-individual variance. Fiber intake increased consistently when MED meals were delivered, with gains observed at breakfast (+2 g), lunch (+2 g), and snacks (+4 g), while sodium intake declined at both lunch (−500 mg) and dinner (−1,200 mg) (**Fig. 2b)**. At the macronutrient level, MED diet adherence was associated with significantly lower intake of carbohydrates, total sugars, and saturated fatty acids (SFA) (p < 0.001), whereas total fat, protein, monounsaturated fatty acids (MUFA), and polyunsaturated fatty acids (PUFA) did not differ significantly between phases. Consistent with these macronutrient shifts, total energy intake was significantly reduced during the MED diet period (p < 0.001), with inter-individual variability in caloric intake notably attenuated by Day 14 (**Fig. 2c, inset)**. Analysis of food group composition revealed that the MED diet was characterized by significantly lower consumption of beverages (p < 0.001), dairy and eggs (p < 0.001), sweets (p < 0.001), and fats and oils (p < 0.001) relative to the habitual diet (**Fig. 2d, S3**). Fruits and juices were marginally reduced (p < 0.05), while baked product intake did not differ significantly between phases. Together, these findings are consistent with a shift toward a lower-calorie, lower-sugar, higher-fiber profile; estimates are model-derived without a cohort-matched gold standard.

**Figure 2:**
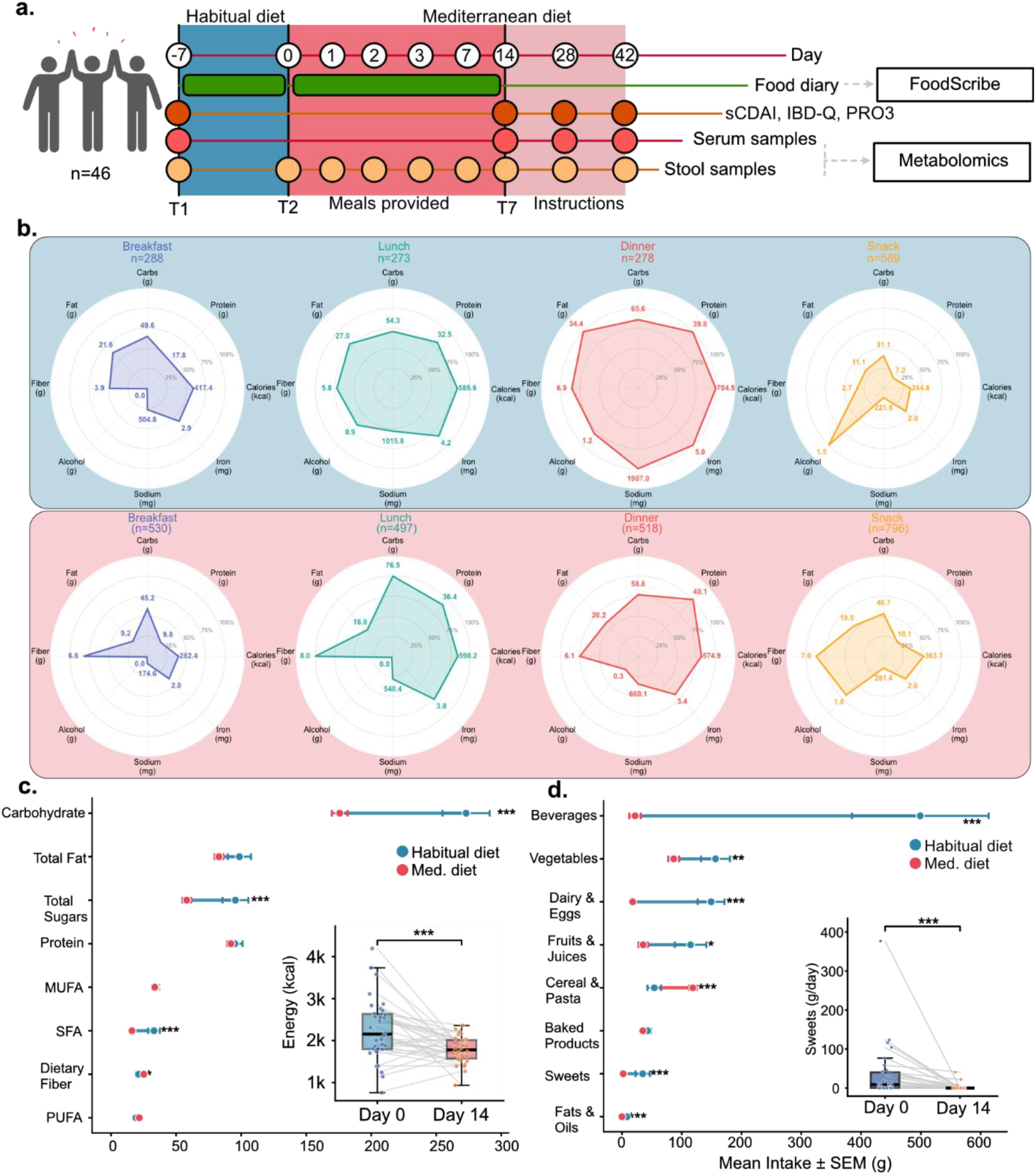
Mediterranean dietary interventional trial: (**a**) Schematic overview of study design (n=46) with dietary intake data collected through first 14-days, clinical assessments (SCDAI, IBD-Q, PRO3) performed, serum samples, and stool samples were collected at indicated timepoints. (**b**) Radar plot for macro and select micronutrient composition in participant meal habits between T1 and T7 timepoints. (**c-d**) Mean macronutrient and food group intake (± SEM) comparing habitual diet (blue,) and Mediterranean diet (red) phases.

Using collected stool samples during the study, we collected mass-spectrometry based metabolomics data to understand the relationship between dietary habits and fecal metabolome^20^. Exploratory, unadjusted correlation anlaysis suggested that fiber and vegetable intake were most strongly associated with a fecal metabolite cluster (**Fig. 3**), including biogenic amines (tyramine, phenylethylamine, GABA) and phenolic acid catabolites (cinnamic acid, p-coumaric acid), with no comparable associations in plasma (**Fig S5**). The stool-specific pattern across biogenic amines and phenolic catabolites is consistent with fermentative processing of fiber-rich foods. Conversely, fruit and juice intake showed negative associations with this metabolite cluster, potentially from the sugar-associated shifts away from proteolytic and phenolic acid-metabolism^21^. Analysis of habitual dietary intake from timepoint T1 to T2 revealed an inverse association between pantothenic acid consumption and disease activity indices (sCDAI, PRO-2, PRO-3, and liquid stool frequency), indicating that patients with more clinical symptoms reported lower pantothenic acid intake (**Fig S4**).

**Figure 3:**
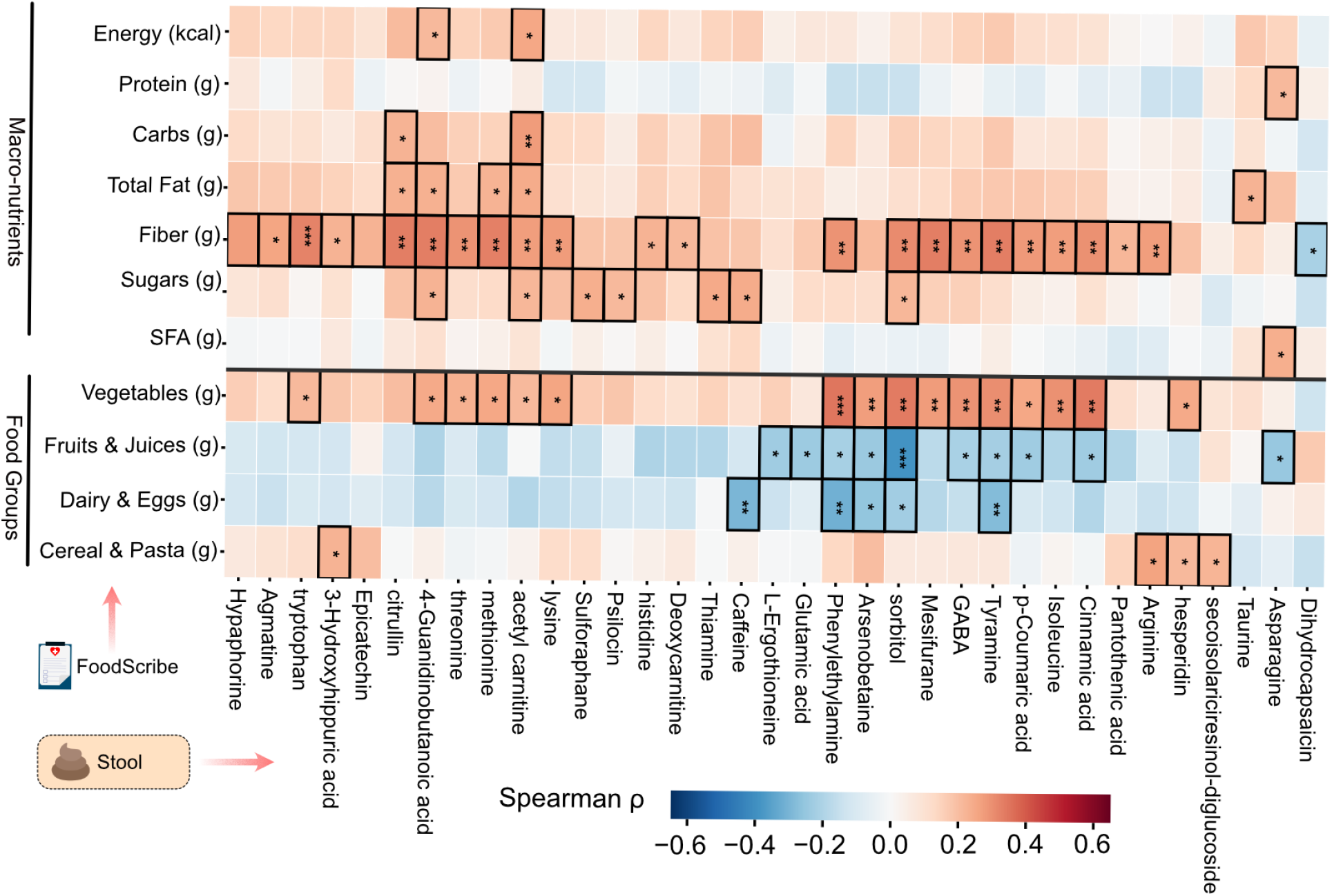
Fecal metabolomics with dietary records predicts response. Spearman correlation heatmap between fecal dietary metabolites (n=90 samples) with macronutrient intake and food groups.

Several limitations warrant consideration. First, FoodScribe’s nutrient inference for ambiguous or culturally specific food descriptions remains imperfect, as entries that lack sufficient specificity for reliable parsing, such as “had a quick bite at a friend’s BBQ” are appropriately returned as low-confidence outputs. Three tested LLM models consistently performed poorly on the descriptions based on the countries (**Fig S1**). Second, gram quantity estimation for foods reported without explicit portion information (e.g., “some rice”) is probabilistic and introduces uncertainty independent of the semantic matching accuracy. Third, LLM outputs are sensitive to model version; all results reported here are tied to specific versioned model strings (claude-sonnet-4-6, deepseek-chat V3.2, gpt-5-mini) and should be re-benchmarked if updated model versions are used. Finally, the metabolomics associations reported here are correlational and do not support mechanistic conclusions; large, prospective and interventional studies are required to establish causal diet-disease relationships.

While FoodScribe enhances existing dietary studies with speed and scalability for analysis, its effectiveness depends on both the quality of food database entries and semantic matching, which may occasionally result in mismatches where expert oversight could be beneficial, while substantially reducing the time required for large-scale dietary annotation compared with manual review. FoodScribe can be adapted to any available custom food databases and will need to be benchmarked for accuracy with new entries added. The transparent and interpretable framework of FoodScribe can facilitate cross-cohort level dietary analysis, for population level studies, by providing harmonized comprehensive nutrient data for integration with other -omics information.

## Data Availability

The data supporting the findings of this study are included within the article and its Supplementary Information files. The foodscribe is available as a tool in the Github repository (https://github.com/harsha-gouda/foodscribe) and the raw metabolomics data for the CCFA are deposited in public metabolomics data repository MassIVE (MSV000098242).

## Acknowledgment

This project is supported by Crohn’s and Colitis Foundation (CCF, 1243263 & 670398), Helmsley Foundation and National Institute of Diabetes and Digestive and Kidney Disease (R01-DK136117) granted to P.C.D. H.G. acknowledges support from Eric & Wendy Schmidt AI in Science Postdoctoral Fellowship.

## Author Contributions

Conceptualization, H.G., P.C.D.; methodology, H.G.,; formal analysis, H.G., J.A., M.C., S.P.G., A.N., investigation, H.G., T.H., B.B., M.G., P.C.D, ; writing – original draft, P.C.D., and H.G.; writing – review & editing, all authors; supervision, P.C.D., funding acquisition, P.C.D. The use of AI in preparation of this manuscript: In the Dorrestein Lab, the use of AI, generative AI, and large language models (LLMs), and software that uses these technologies, both free and commercial, is encouraged across all aspects of research, including literature review, coding, data analysis, and text editing.

## Notes

The authors declare the following competing financial interest(s): P.C.D. is an advisor and holds equity in Cybele, Sirenas, and BileOmix, and he is a scientific co-founder, advisor, and holds equity to Ometa, Enveda, and Arome with prior approval by UC San Diego. P.C.D. consulted for DSM Animal Health in 2023. B.S.B has received consulting fees from Janssen, Sanofi, Merck, Abbvie, Celltrion and has received research grants from Merck, Mirador, Gilead, S.R.T. Therapeutics. M.G. has received research grants from AbbVie and Janssen.

## Ethics Declaration

The Mediterranean dietary intervention study was conducted at the University of California San Diego and was approved by the Institutional Review Board of the University of California San Diego (IRB #800805). All participants provided written informed consent before enrolment. The study was conducted in accordance with the Helsinki Declaration of 1975, as revised in 2000. The trial was registered on ClinicalTrials.gov as Effect of Mediterranean Diet in Inflammatory Bowel Disease (NCT05973500).

## Experimental and analytical methods

### Meal Information retrieval

A multi-step computational pipeline was adopted to convert free text food dairy entries into structured nutrient profiles, based on nutrient labels available in the USDA Foodcentral database. To parse meal ingredients from the free text meal description, we used an existing Large language model (Claude/GPT/deepseek) to parse the text, and return a structured JSON list of ingredients with estimated quantities(grams), units and confidence scores. The output JSON contained a list of objects {Ingredients, qty, unit, grams, confidence} for each meal description that passed the query. Final prompt after multiple rounds of optimization for accuracy.

<QUERY>:

You are a helpful assistant that extracts structured information from short meal descriptions. Return an empty array [] if no food items can be identified.

Given a single meal text, output ONLY a JSON objects for each food item with these fields:

- Ingredient: canonical short item name (string) that closely matches to the food names in FoodData Central
- qty: number if explicit, else null
- unit: unit string if explicit (e.g., ‘cup’,’piece’,’g’), else null
- grams: number if you can directly infer grams, where the exact quantity cannot be determined, provide a reasonable estimate instead of leaving it unspecified.
- confidence: give a score in the range of 1 to 5 with 5 being high confidence Return only JSON and nothing else.

Meal description: {*meal_description*}

### Nutrient retrieval

To retrieve nutrients for each meal, ingredients retrieved from the large language model was used to query the available USDA FoodCentral database. We converted the food descriptions into semantic embeddinging using sentence transformers from the MPNet model built on top of microsoft/mpnet-base (text-embedding-3-large, 3072 dim). Cosine similarity search is run against a pre-built index of 13,590 USDA food descriptions(Foundational, legacy and survey foods), and 112,683 food descriptions from the NHANES survey (2921-2023) to find the best-matching USDA food entry. We observed that using the USDA database of branded foods reduced the accuracy of prediction and semantic retrieval, and hence the foods only from the Foundational, SR Legacy and FNDDS (Survey foods) were encoded using the sentence transformer to create the embeddings. Based on the text description with context from the meal, the nearest neighbour fdc_id was retrieved that closely resembles the ingredient description based on cosine similarity as a distance metric. At this stage, for each ingredient, fdc_id was retrieved that was then mapped to nutrients from the USDA database. The nutrient profiles were adjusted based on the amount/volume of ingredients in the meal retrieved by the large language model. Nutrients are summed across ingredients to produce meal-level totals. Macronutrient energy split (% protein, carb, fat) is calculated. Results are displayed as a formatted table or exported to CSV.

The fdc_id lookup directly from the LLM prompting was attempted, however had higher rates of false positives and negatives, from retrieval of non-existing fdc_id possibly due to hallucinations. We also have added additional features to create embeddings based on user needs, to train models, if the nutrient profiles are available in their costume database using sentence transformer embedding model (text-embedding-3-large, 3072 dim).

### Cost associated with FoodScribe analysis

Based on the models used in the manuscript the claude-sonnet-4-6 ($2.35) was more expensive compared to gpt-5-mini ($1.81) and deepseek-chat ($0.10) for analysis of 1000 meal descriptions.

### Dietary journal data collection

A 7-day habitual diet was collected using the diet journal as a part of Mediterranean dietary interventional trial at the University of California San Diego. Ethical approval for the study was granted by the Institutional Review Board (IRB) at University of California at San Diego. Healthy individuals and people with irritable bowel syndrome were asked to complete a daily diet log for seven days before starting the diet and 14 days during the course of the Mediterranean diet. A total of 3,807 free text meal descriptions were obtained from 50 individuals that were analyzed using the foodscribe analysis pipeline.

### Human Dietary Data Processing and Nutrient Estimation

Dietary intake data were collected via participant-reported food journals and subsequently reviewed and annotated by a qualified nutritionist (M.S.C.). Nutritional composition for each food item was primarily derived from the United States Department of Agriculture (USDA) FoodData Central database. For food items of Spanish origin or those not available in the USDA database, nutritional values were obtained from the Base de Datos Española de Composición de Alimentos (BEDCA). When participants reported quantities using standardized volumetric measures (e.g., ¾ cup, 1 tablespoon), the corresponding weight or volume was applied directly as specified by each database entry. For food items reported without precise measures (e.g., “one banana,” “a handful of nuts”), standard reference weights were assigned based on commonly accepted portion sizes (e.g., one medium banana ≈ 110 g, one serving of nuts ≈ 30 g). For each meal entry, total energy (kcal), carbohydrates (g), protein (g), fat (g), and dietary fiber (g) were calculated by summing the individual contributions of each food component within that entry.

### Clinical data acquisition

The study was approved by the Institutional Review Board of the University of California San Diego (IRB #800805). Participants attended three study visits throughout the intervention period. At each visit, clinical assessments were performed, biological samples were collected. During the baseline visit, participants underwent initial evaluations to establish pre-intervention measures. At the second visit, participants received detailed instructions on how to follow the anti-inflammatory dietary intervention. They were instructed to adhere strictly to the prescribed diet for the subsequent two weeks. To facilitate compliance, participants were provided with pre-prepared meals delivered to their homes for lunches and dinners, as well as groceries and ingredients needed to prepare the recommended breakfasts and snacks according to the dietary protocol. At the final visit, conducted approximately six weeks after initiation of the dietary intervention, adherence to the intervention, participant satisfaction, and changes in clinical outcomes were evaluated. During each visit, IBD patients were asked to complete an sCDAI and IBD-Q questionnaires, recording symptoms experienced during the 7 days prior to the visit. Diet logs were recorded between visits and were to be brought, upon completion, to the next appointment.

Stool samples were collected at each study visit using sealed collection kits provided by the research staff. Participants collected the samples at home using nylon-flocked swabs, which were immediately transferred into 1 mL Matrix Tubes (Thermo Fisher Scientific, catalog #3741) containing 400 µL of 95% ethanol, following the extraction protocol described by Brennan et al. (2024). This approach was designed to minimize well-to-well contamination while preserving samples for downstream microbiome and metabolomic analyses. Samples were transported in temperature-controlled containers, aliquoted upon arrival, and stored at −80 °C until further processing.

### Metabolomics data analysis

Stool and plasma samples were collected from 32 adult volunteers, including healthy individuals and participants diagnosed with Crohn’s disease (CD), before and after a 14 day Mediterranean diet intervention under approved protocols from the University of California San Diego (IRB #800805). Sample preparation was performed following a previous study protocol (Anal. Chem. 2026, 98, 4, 3160–3176). For each sample, 20 μg of stool extract was aliquoted into centrifuge tubes containing a steel bead followed by the addition of 800 μL of ice-cold (4 °C) extraction solvent (50:50 methanol/water, LC–MS grade) spiked with 500 nM sulfadimethoxine as an internal standard to track injection and instrument behaviors at each well level. Briefly, after homogenization and centrifugation, 150 μL of the resulting supernatant was transferred into shallow 96-well plates and dried using a vacuum centrifuge concentrator (room temperature, ∼3 h). The dried extracts were reconstituted in 150 μL of 50:50 methanol/water and centrifuged at 2000*g* for 10 min at 4 °C, and 100 μL of the clarified supernatant was transferred to fresh plates and stored at −80 °C until LC–MS/MS analysis.

## Statistical analysis

### Accuracy Metrics

For each nutrient– LLM model pair, mean Absolute Error (MAE) was calculated as the average of the absolute differences between ground truth and estimated values. Root Mean Squared Error (RMSE) was calculated as the square root of the mean squared differences, penalising larger errors more heavily than MAE.

### Performance evaluation

To evaluate performance of the foodscribe analysis pipeline, nutribench nutrient profiles were compared against the foodscribe outputs, to determine MAE and calculated F-statistics with defined (50, 100, 150, 200 kcal) threshold from the nutribench estimates. Accuracy metrics were also obtained for the same model (sonnet4.6) by repeated analysis, by comparing output from the same reasoning model.

### Longitudinal dietary record analysis

To assess longitudinal changes in dietary intake over the intervention period, macronutrient and micronutrient intake at baseline (T1) and follow-up (T7) were compared in subjects with paired observations at both timepoints (n=46). For each nutrient, a Wilcoxon signed-rank test was applied to the paired T1 and T7 values, with individual subject trajectories visualized as connecting lines between timepoints. Statistical significance was denoted as * p<0.05, ** p<0.01, and *** p<0.001.

### Dietary record data integration with metabolomics

To examine associations between fecal metabolite abundance and dietary intake, Dietary data from timepoints T2 through T7 were merged with corresponding fecal metabolomic data by subject and timepoint. Food group intake (gram weight) was derived from dietary records across four categories: vegetables and vegetable products, fruits and juices, dairy and egg products, and cereal grains and pasta. Spearman correlations were computed between each metabolite and each dietary variable (macronutrients and food groups), requiring a minimum of six paired observations and excluding diet columns where more than 85% of values were zero (to avoid spurious correlations driven by non-consumption). Metabolites with at least one significant association (p<0.05) were retained. Molecule rows were ordered by Ward hierarchical clustering. Associations between plasma metabolite levels at follow-up (V2) and concurrent dietary intake at T7 were evaluated using the same Spearman correlation framework. Plasma metabolite peak areas were averaged across technical replicates per subject and visit. Dietary variables included seven macronutrients and four food group categories.

## Supplementary Figures

**Figure S1:**
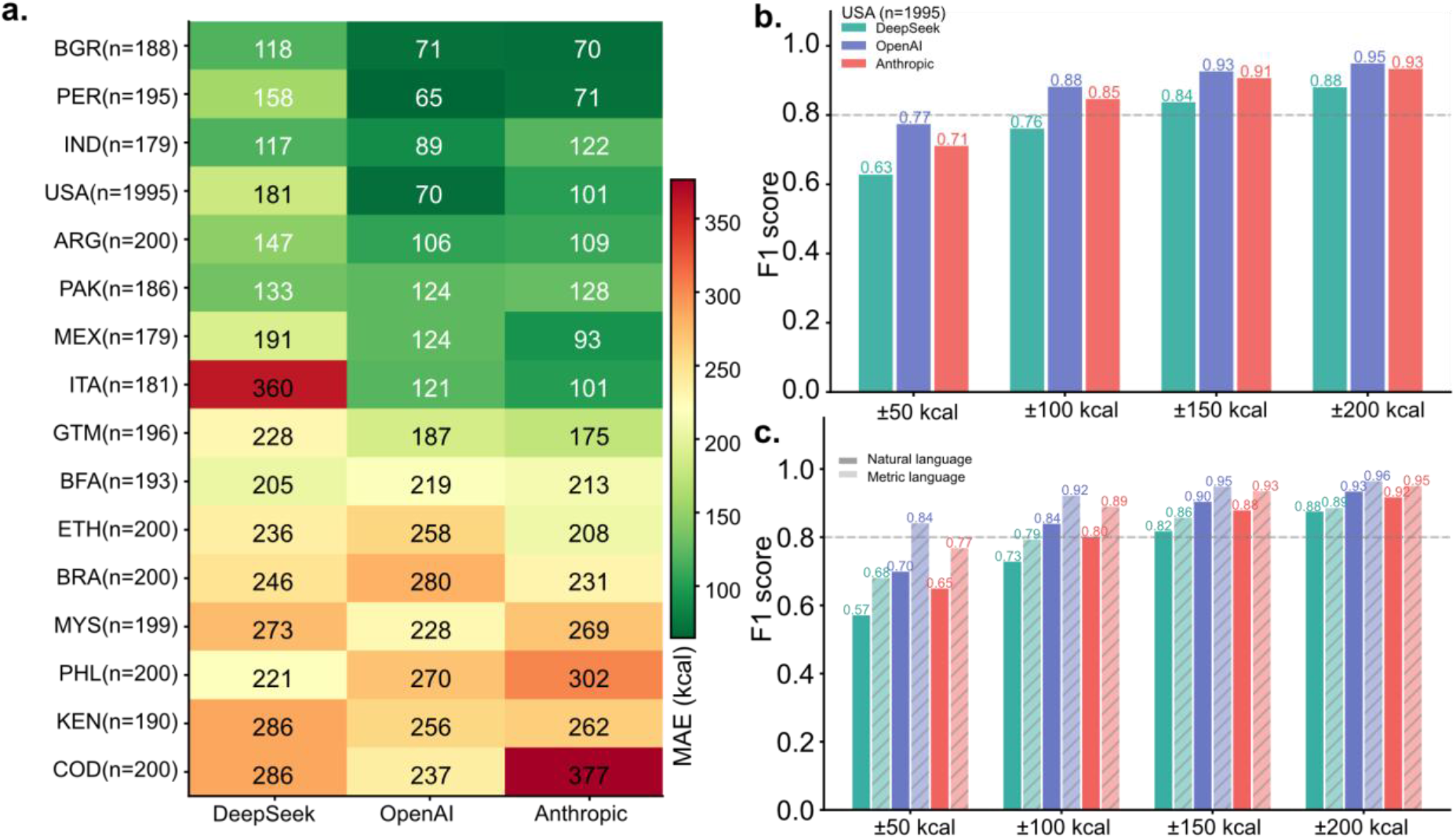
Evaluation of LLM-based nutrient estimation using the Nutribench dataset. (a) Mean absolute error for caloric estimation (kcal) of meal description from 16 countries using three-LLM providers, NutriBench curated ground truth values (n=4,881 meals).Countries are ordered by ascending average MAE across models. Cell color represents MAE magnitude (green = lower, red = higher). Sample sizes per country are indicated in parentheses. (b) Tolerance-based F1 scores for caloric estimation in US-based meals (n=1,995) at four absolute tolerance thresholds (±50, ±100, ±150, and ±200 kcal). A prediction was classified as a true positive if it fell within the specified tolerance of the curated ground truth value; false positives and false negatives represent systematic overestimates and underestimates beyond tolerance, respectively. (c) Tolerance-based F1 scores for caloric estimation in US-based meals stratified by serving description format: natural language (solid bars) versus metric/gram-based descriptions (hatched bars), at the same four tolerance thresholds as in (b). Metric descriptions consistently yielded higher F1 scores across all models and thresholds.

**Figure S2:**
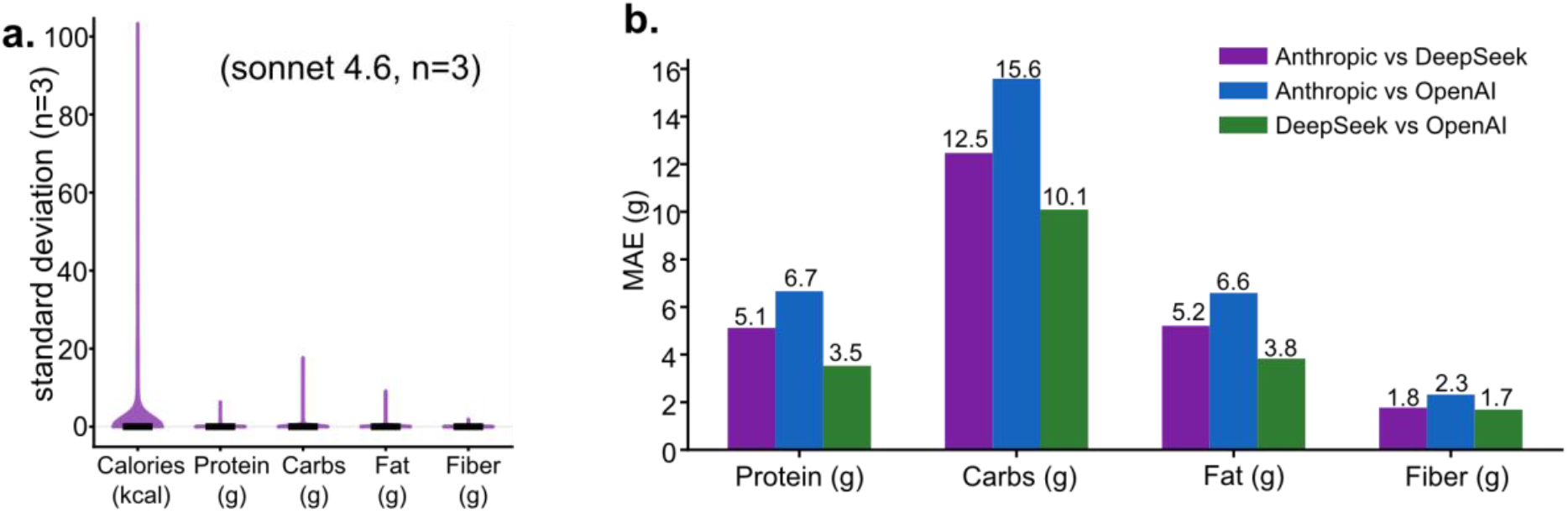
Inter-model agreement in nutrient estimation in MED dietary records. (a) Model reproducibility of macronutrient estimates for claude-sonnet-4.6 across three independent runs (n=3), demonstrates high run-to-run consistency. (b) Pairwise means absolute error (MAE) between LLM providers (Anthropic vs. DeepSeek, purple; Anthropic vs. OpenAI, blue; DeepSeek vs. OpenAI, green), suggesting that defined nutrients are estimated more reliably across models.

**Figure S3:**
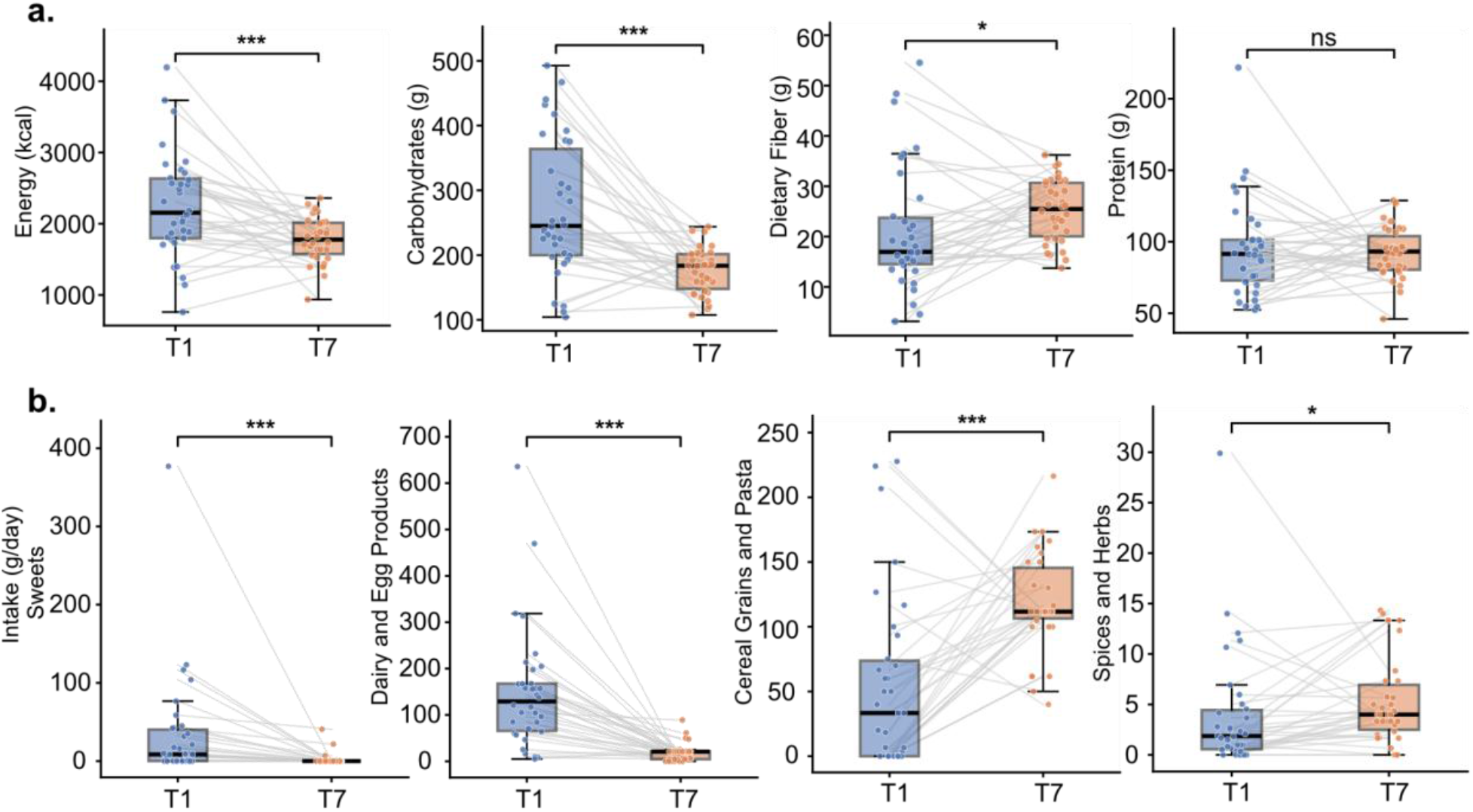
FoodScribe-derived macronutrient and food group changes between habitual (T1) and Mediterranean diet (T7) phases in a dietary intervention cohort. (a) Paired boxplots comparing macronutrient intake at T1 (habitual diet, blue) and T7 (Mediterranean diet, orange) across 52 participants. Lines connecting paired observations illustrate individual-level trajectories, highlighting substantial inter-individual variability in the magnitude of dietary change. **(b)** Paired boxplots of food group intake (g/day) between T1 and T7 for selected categories. Statistical significance was assessed by Wilcoxon signed-rank test; *p < 0.05, ***p < 0.001.

**Figure S4:**
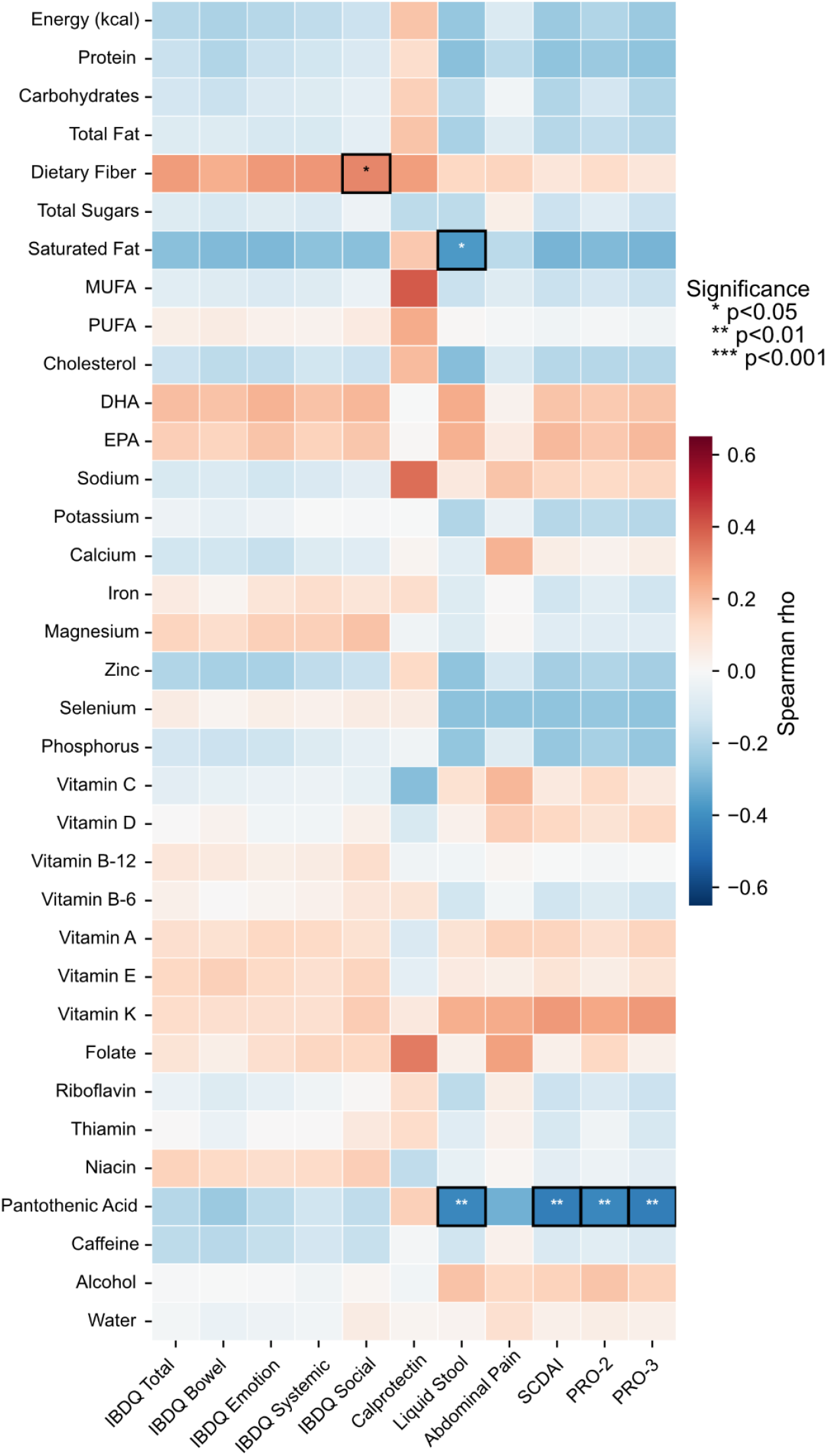
Spearman Correlation Between Baseline Dietary Intake and Clinical Disease Indices. Heatmap depicting Spearman correlation coefficients (ρ) between 35 dietary nutrients and 11 clinical indices measured at baseline (V1) in IBD patients and controls. Nutrients (rows) include macronutrients, fatty acids, vitamins, minerals, and other dietary components derived from 3-day averaged dietary records. Clinical indices (columns) include IBDQ subscores (Total, Bowel, Emotion, Systemic, Social), fecal calprotectin, liquid stool frequency, abdominal pain, SCDAI, PRO-2, and PRO-3. Cell color reflects the Spearman ρ value (red = positive correlation, blue = negative correlation; scale clamped at ±0.65). Asterisks denote statistically significant correlations (* p<0.05, ** p<0.01, *** p<0.001); no correction for multiple comparisons was applied.

**Figure S5:**
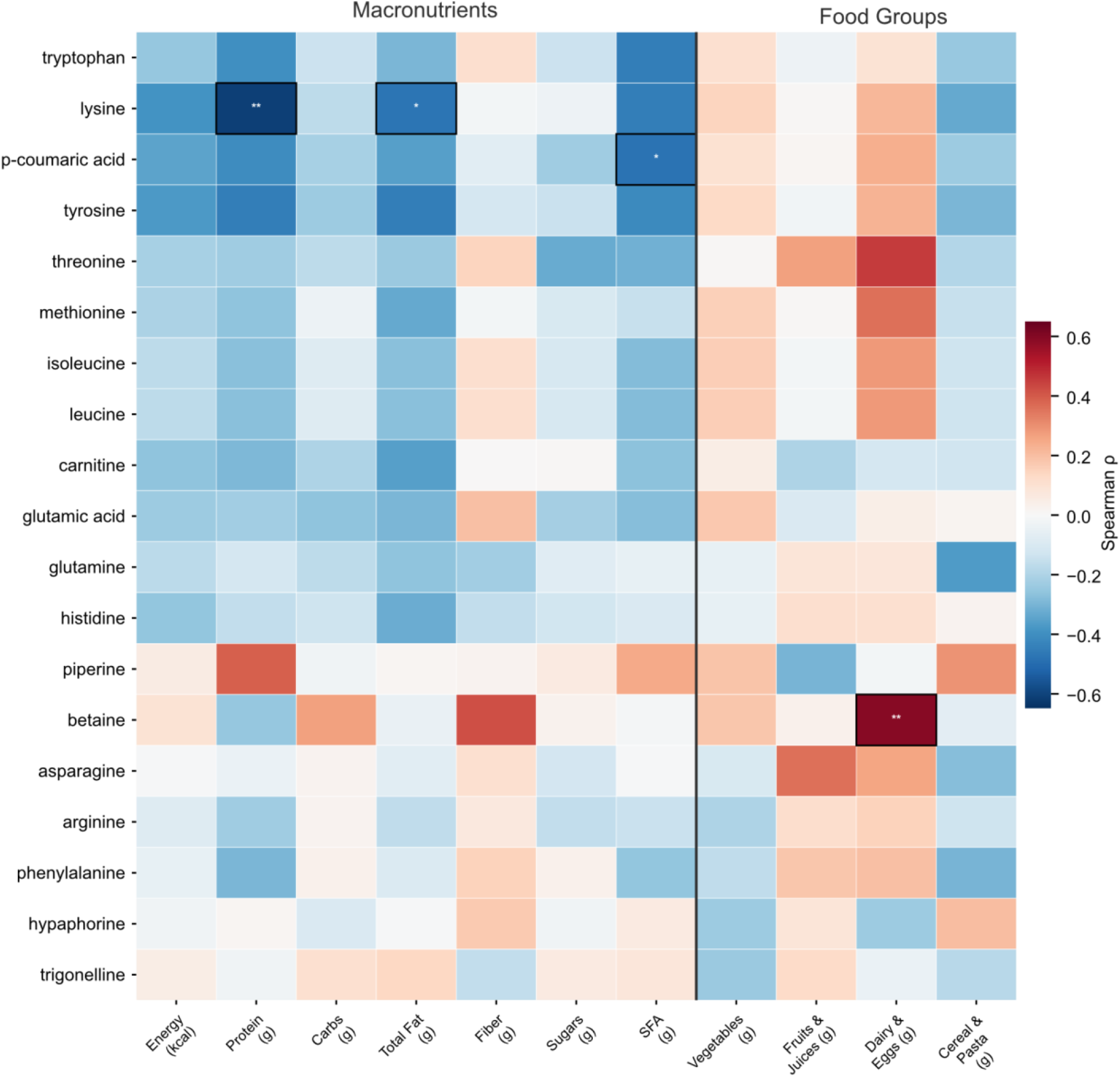
Association Between Plasma Metabolites and Dietary Intake. Heatmap displaying Spearman correlation coefficients (ρ) between 19 plasma metabolites measured at follow-up (V2) and dietary intake at the corresponding timepoint (T7) in n=19 subjects. Plasma metabolite levels (rows) were derived from targeted mass spectrometry and represent mean peak area across technical replicates. Cell color reflects the Spearman ρ (red = positive, blue = negative; scale clamped at ±0.65). Cells with p<0.05 are outlined in black and annotated with asterisks (* p<0.05, ** p<0.01, *** p<0.001); no correction for multiple comparisons was applied.

